# Efficacy of Tenapanor in Managing Hyperphosphatemia and Constipation in Hemodialysis Patients: A Randomized Controlled Trial

**DOI:** 10.1101/2025.02.02.25321552

**Authors:** Naoki Suzuki, Yuuki Takeda, Akie Kabuto, Takao Konishi, Takahiro Konishi, Fumi Sumino, Hayanari Iwata, Mone Iwagami, Yusuke Kouchi, Yasumasa Hitomi, Toru Takatani, Masato Nishimura, Nodoka Sato

## Abstract

**Introduction:** Tenapanor is a minimally absorbed, small-molecule inhibitor of sodium/hydrogen exchanger 3 and thus suppresses sodium absorption in the gastrointestinal tract. It is approved by the FDA for the treatment of hyperphosphatemia in dialysis patients. This randomized controlled trial evaluated its efficacy in the treatment of hyperphosphatemia and constipation in hemodialysis patients.

**Methods:** Ninety hemodialysis patients were randomized 1:1 to receive either tenapanor or standard care. Randomization was performed using a computer-generated sequence stratified by baseline serum phosphorus levels. The tenapanor group began treatment with a dosage of 10 mg/day, which was adjusted based on serum phosphorus levels. Primary outcomes were changes in serum phosphorus levels in the tenapanor and control groups and changes in stool consistency, assessed weekly using the Bristol Stool Form Scale (BSFS) in the tenapanor group. Secondary outcomes included laxative use and phosphate binder prescription patterns. Serum phosphorus levels, serum calcium, albumin, and related biochemical parameters were monitored every two weeks. Data were analyzed using intention-to-treat principles. This study was not blinded.

**Results:** Of the 90 randomized participants, 69 completed the 23-week study. Tenapanor significantly improved stool consistency and resolved constipation (BSFS types 1-2) by week 5. A transient increase in loose stools (BSFS types 6-7) occurred early, with 10 participants discontinuing due to diarrhea. Laxative use decreased significantly in the tenapanor group, from 58.2% at baseline to 35.6% at week 23 (p < 0.01). Serum phosphorus levels were decreased in both groups, with comparable control. Lanthanum carbonate prescriptions decreased significantly in the tenapanor group and were largely replaced by low-dose tenapanor.

**Conclusion:** Tenapanor improves stool consistency, reduces laxative use, and provides effective phosphorus control in hemodialysis patients and represents a promising alternative to conventional phosphate binders.

**Funding:** None.

**Trial Registration:** Registered with UMIN-CTR (UMIN000033778; http://www.umin.ac.jp/ctr/index.htm).

## Introduction

Hyperphosphatemia has been independently associated with coronary and aortic calcification, as well as cardiovascular and all-cause mortality in end-stage renal disease[1-5]. Therefore, the management of hyperphosphatemia is an important part of the routine care of patients on chronic dialysis[6]. Currently used phosphate binders include calcium-based options such as calcium carbonate and non-calcium-based options such as sevelamer and lanthanum carbonate[7]. However, these phosphate binders can cause gastrointestinal side effects and elevate medication burden, which can lead to poor patient adherence and reduce treatment efficacy[8-10]. In addition, there are concerns about the long-term safety of lanthanum carbonate, which has been reported to form lanthanum deposits in the stomach and duodenum[11,12].

Tenapanor is a minimally absorbed, small molecule inhibitor that targets sodium/hydrogen exchanger 3 (NHE3) and thus reduces sodium absorption in the gastrointestinal tract. It was initially highlighted as a new treatment for constipation-predominant irritable bowel syndrome[13], but has since also shown promise in the treatment of hyperphosphatemia due to its ability to reduce passive phosphate absorption in the intestinal tract[14]. On October 17, 2023, the FDA approved tenapanor as the first phosphate binder adjunctive therapy to lower serum phosphorus levels in adults on dialysis. Previous studies have reported that gastrointestinal adverse events were common with tenapanor, with the most common adverse event, diarrhea (mostly mild diarrhea), occurring in 63.7% to 74.4% of patients[15-18]. As tenapanor inhibits sodium absorption, this effect is thought to be mainly due to the enhancement of colonic propulsive activity by osmotic pressure-induced increase in intestinal content[19]. We hypothesized that long-term use of tenapanor could normalize the stools of dialysis patients and reduce the amount of laxative used. In this study, we conducted a randomized controlled trial to test this hypothesis and to clarify the effect of tenapanor on serum phosphorus levels and stool consistency in actual clinical practice.

## Materials and Methods

### Participants

This parallel-group, randomized, controlled trial was conducted to evaluate the effects of tenapanor administration on serum phosphorus control, stool consistency, and number of laxative prescriptions. Participants were recruited between February 1, 2024, and February 28, 2024, from patients at the clinic affiliated with Tojinkai Hospital. Patients were excluded (1) who had not been undergoing hemodialysis for at least 2 years at baseline, (2) who had a history of or current inflammatory bowel disease or diarrhea-type irritable bowel syndrome, and (3) whose C-reactive protein (CRP) was 1.0 mg/dL or higher. Clinical staff screened for eligibility and enrolled participants. Participants who did not meet these exclusion criteria were randomized 1:1 to receive either tenapanor or standard care using a computer-generated sequence stratified by baseline serum phosphorus level (≤5.5 mg/dL or >5.5 mg/dL), with allocation concealment ensured by a secure computerized system. There were no restrictions on block size. The study was conducted as an open-label study, and no changes were made to the study methods, including eligibility criteria, data collection procedures, or statistical analysis plans after the study was initiated.

Sample size was calculated based on the primary outcome of changes in stool consistency as assessed by the Bristol Stool Form Scale (BSFS). Assuming a mean difference of 1.0 BSFS units between the tenapanor and control groups, with a standard deviation of 1.2 BSFS units[16], 90% power, and a two-sided significance level of 5%, a minimum of 31 participants per group was required. To account for an estimated 15% dropout rate, the target sample size was set at 90 participants (45 per group).

This study was conducted in accordance with the principles of the Declaration of Helsinki and adhered to all applicable ethical guidelines. Ethical approval was granted by the Ethics Committee for Human Research at Tojinkai Hospital (Approval Number: 2024-03). Written informed consent was obtained from all participants before their enrollment in the study. Participants were provided with detailed information about the research objectives, procedures, their right to withdraw at any time without consequence, and the measures in place to ensure the confidentiality and anonymity of their data. The trial was retrospectively registered with the University Hospital Medical Information Network Clinical Trials Registry under trial number UMIN000033778 (UMIN-CTR URL: http://www.umin.ac.jp/ctr/index.htm) on 25 December 2024 due to an administrative oversight.

### Data Collection

The observation period for this study was March 11, 2024, through August 24, 2024. In the tenapanor group, tenapanor administration started on March 11, 2024, and the start date of administration. was considered the baseline. The initial dose was 10 mg/day administered orally, and the dose was adjusted every two weeks based on serum phosphorus levels. Dose adjustments were made at the discretion of the treating physician to optimize phosphorus control. Participants in the control group did not receive tenapanor but continued their existing laxative regimen. Commonly prescribed laxatives included lanthanum carbonate hydrate, precipitated calcium carbonate, and Iron-based phosphate binder, with dosages based on participants’ prior use. No changes were made to laxative prescriptions unless clinically indicated. Stool consistency were assessed weekly from baseline through week 7 by patient interview using the BSFS[21]. Blood samples (10 mL) were collected at the time of hemodialysis for measurement of routine biochemical and hematological factors, including hemoglobin, urea nitrogen, creatinine, calcium, inorganic phosphorus, sodium, albumin, β2-microglobulin (β2-MG), and CRP. Single pool Kt/Vurea was calculated using the formula of Shinzato et al. to evaluate dialysis efficiency. The proportion of each prescribed phosphate binder and laxative was calculated by dividing the number of prescriptions for that prescription medication by the total number of prescriptions for all phosphate binders and laxatives combined. Blood samples were collected at the beginning of the week (Monday or Tuesday) and analyzed for other key biomarkers every two weeks.

### Statistical Analysis

Baseline characteristics were compared using the Welch *t*-test for continuous variables and the chi-square test for categorical variables. Changes in blood data between the tenapanor and control groups were analyzed using repeated measures analysis of variance. Changes in variables (proportion of phosphate binder prescriptions, BSFS score, and proportion of laxative prescriptions) within the tenapanor group were visualized using a Sankey diagram. Changes in laxative prescriptions at baseline, week 7, and week 23 were evaluated using the McNemar test with Bonferroni correction to adjust for multiple comparisons. Normally distributed values are reported as mean ± standard deviation, while non-normally distributed values are presented as median with interquartile range. Cohen’s d was used to calculate the standardized effect size of the differences in blood data at week 23 between the tenapanor and control groups. Data were analyzed using intention-to-treat (ITT) principles, and a p-value < 0.05 was considered statistically significant. All analyses were conducted using R software (version 4.0.3). No interim analyses or pre-specified stopping guidelines were planned for this study.

## Results

A total of 136 adult patients were enrolled in the study. The process by which patients were enrolled is shown in Figure 1. Of all the patients, 46 met one or more of the exclusion criteria, and 90 were randomized into the study. During the observation period, in the tenapanor group, the study was discontinued due to diarrhea symptoms or frequent bowel movements in 10 patients and hospitalization or transfer in 4 patients. In the control group, the study was discontinued due to hospitalization, transfer, or death in 7 patients. For the primary outcome, 31 participants in the tenapanor group and 38 participants in the control group were included in the final analysis. All analyses were based on the original group assignments (ITT analysis). The same denominators were used for secondary outcomes, unless otherwise specified. The study was completed as planned, with follow-up concluding on August 24, 2024.

**Figure 1.**
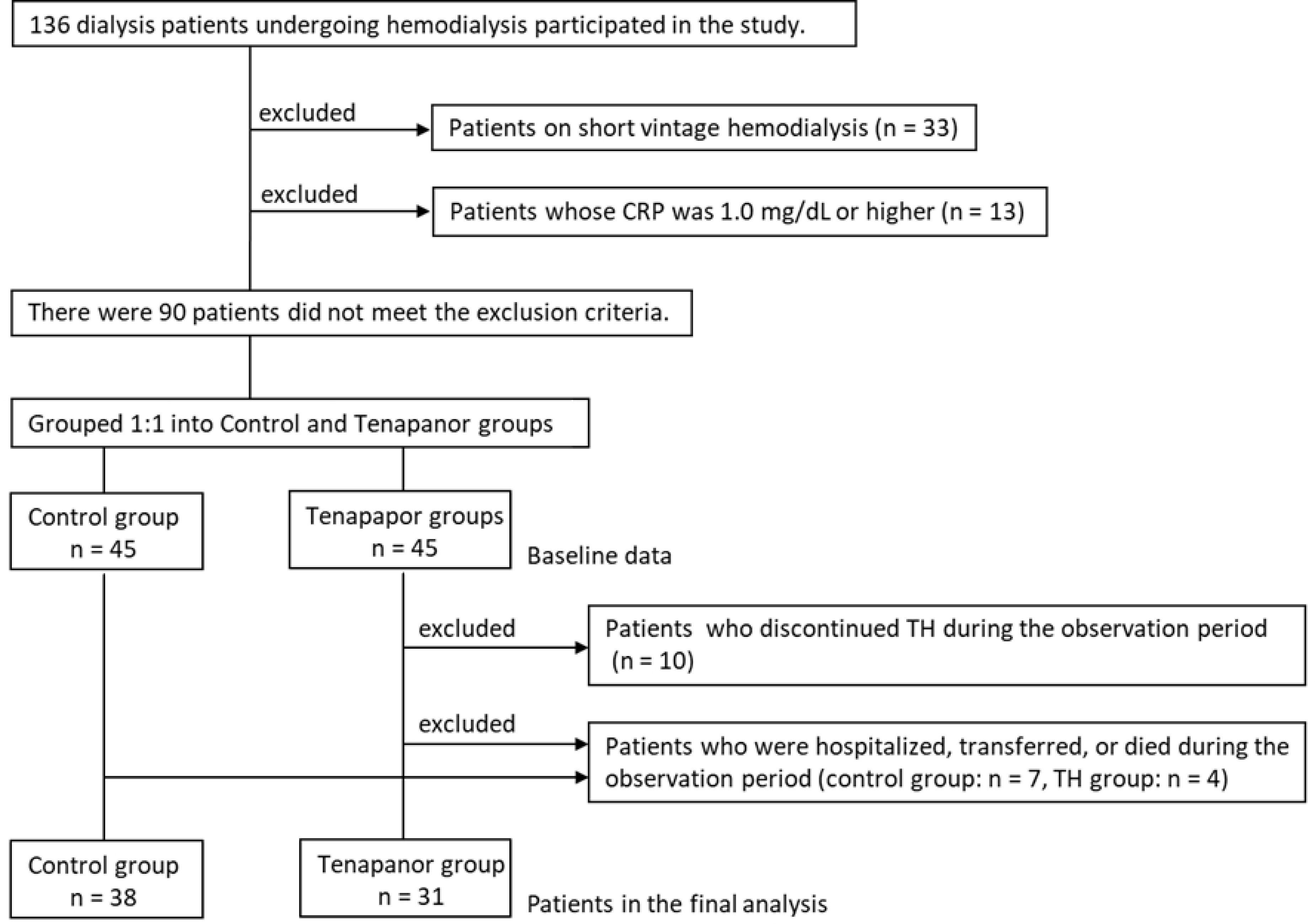
**Flowchart of the study enrollment process. CRP, C-reactive protein; BSFS, Bristol Stool Form Scale.**

Baseline characteristics are shown in Table 1. For the overall population, the mean age was 72.9 years, 57.8% were men, the median hemodialysis vintage was 12.5 years, and 35.6% had diabetes. There were no significant differences between the tenapanor group and the control group in any of the patient characteristics.

**Table 1.** Characteristics of the study participants.

| Variables | Overall (n = 90) | Tenapanor (n = 45) | Control (n = 45) | p value |
| --- | --- | --- | --- | --- |
| Age, years | 72.9±9.0 | 74.4±8.9 | 71.5±9.1 | 0.130 |
| Male, n (%) | 52(57.8) | 29(64.4) | 23(51.1) | 0.286 |
| Hemodialysis vintage, years | 12.5(2.0-22.8) | 14.0(3.0-24.0) | 9.0(3.0-21.0) | 0.722 |
| Diabetes mellitus, n (%) | 32(35.6) | 17(37.8) | 15(33.3) | 0.826 |
| Dry weight, kg | 56.5±11.4 | 56.4±11.3 | 56.5±11.7 | 0.971 |
| Hemoglobin, g/dL | 11.2±1.1 | 11.3±1.2 | 11.2±1.0 | 0.727 |
| Urea nitrogen, mg/dL | 61.4±13.7 | 60.9±14.1 | 62.0±13.5 | 0.709 |
| Creatinine, mg/dL | 9.56±2.17 | 9.60±2.22 | 9.52±2.13 | 0.861 |
| Calcium, mg/dL | 8.77±0.50 | 8.79±0.53 | 8.75±0.48 | 0.679 |
| Inorganic phosphorus, mg/dL | 5.08±1.15 | 5.08±1.28 | 5.08±1.01 | 0.993 |
| Albumin, g/dL | 3.68±0.28 | 3.67±0.24 | 3.69±0.3 | 0.769 |
| C-reactive protein, mg/dL | 0.09(0.05,0.18) | 0.10(0.04-0.16) | 0.08(0.05-0.20) | 0.701 |
| Intact parathyroid hormone, pg/mL | 101.50(55.50,179.00) | 93.0(55.0-198) | 102.0(57.0-176) | 0.952 |
| Single pool Kt/V urea | 1.80±0.35 | 1.75±0.35 | 1.86±0.36 | 0.151 |

A Sankey diagram of the variation in the proportion of phosphate binder prescriptions is shown in Figure 2. The proportion of prescriptions of specific phosphate binders at baseline was highest for lanthanum carbonate hydrate at 55.6%, followed by precipitated calcium carbonate at 35.6%, iron-based phosphate binders at 31.1%, sevelamer hydrochloride at 17.8%, and Bixalomer at 11.1%, which was the lowest. The most common phosphate binder from which patients switched to tenapanor was lanthanum carbonate hydrate at 40.0%. In addition, the proportion of patients who received tenapanor as their initial treatment was 35.6%. Finally, at week 23, the proportion of prescriptions for tenapanor was 68.9% and that for lanthanum carbonate hydrate decreased to 8.9%. On the other hand, the proportion of prescriptions for Bixalomer increased to 31.1%. The proportion of prescriptions for precipitated calcium carbonate, iron phosphate binders and sevelamer hydrochloride were 31.1%, 15.6% and 11.1%, respectively. In addition, 87.1% of the total dose of tenapanor was administered at a dose of 10 mg/day or less (as shown in Figure 3).

**Figure 2.**
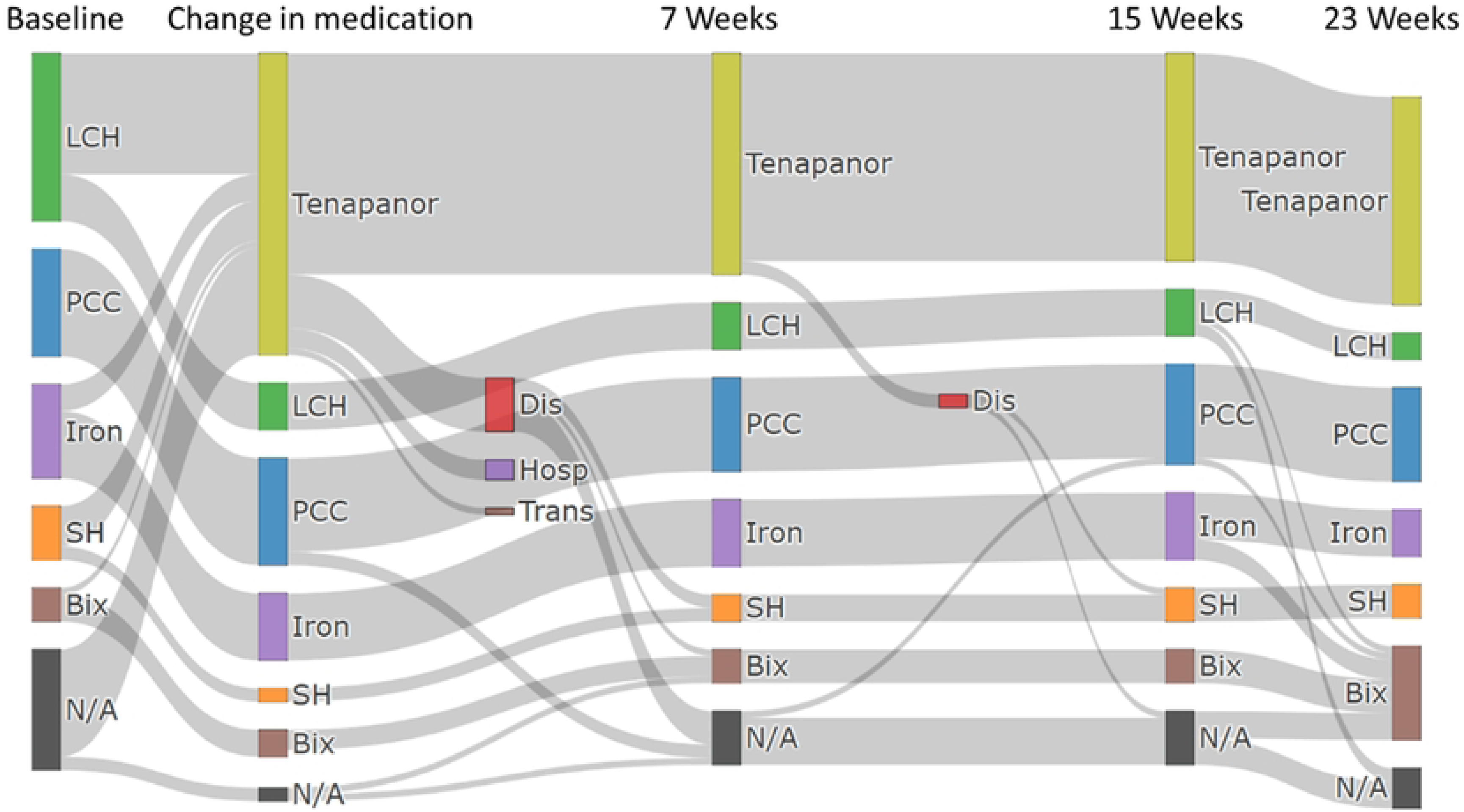
The variations in the prescription of phosphate binders. LCH, lanthanum carbonate hydrate; PCC, precipitated calcium carbonate; Iron, Iron-based phosphate binder; Bix, bixalomer; SH, sevelamer hydrochloride; Dis, discontinuation; Hosp, hospitalization; Trans, transfer to a different hospital.

**Figure 3.**
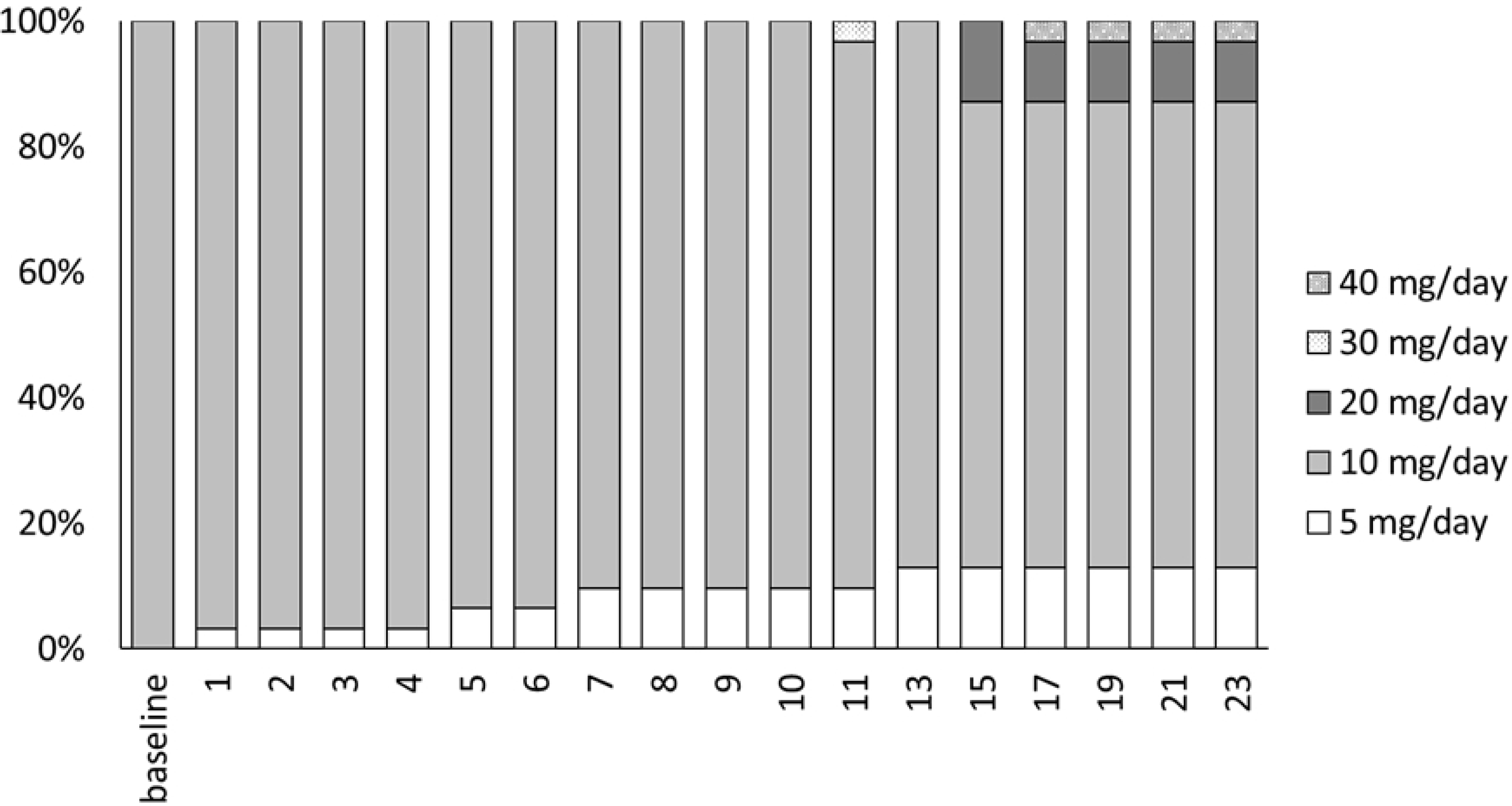
**The proportion of the dosage of tenapanor.**

Changes in serum phosphorus levels in the tenapanor group and the control group are shown in Figure 4. The mean serum phosphorus levels in the tenapanor group and the control groups were 5.17 and 5.06 mg/dL at baseline, 4.59 and 4.99 mg/dL at week 7, and 4.11 and 4.76 mg/dL at week 23 (Cohen’s d = 0.38, 95% confidence interval (CI): -0.05-0.82), respectively. Although serum phosphorus levels tended to decrease significantly in both groups (tenapanor: p < 0.0001, control: p = 0.0029), no significant difference was observed between the two groups (p = 0.1256). Changes in serum-corrected calcium and albumin levels are shown in Supplementary Figure 1 and Supplementary Figure 2. The calcium levels in the tenapanor and control groups were 8.76 and 8.75 mg/dL at baseline, 8.73 and 8.68 mg/dL at week 7, and 8.66 and 8.51 mg/dL at week 23 (Cohen’s d = 0.23, 95%CI: -0.67-0.20), respectively. Although there was no significant change over time in the tenapanor group (p = 0.6270), a significant downward trend was observed in the control group (p = 0.0035). There was no significant difference between the two groups (p = 0.7553). In addition, the albumin levels were 3.65 and 3.68 mg/dL at baseline, 3.59 and 3.59 mg/dL at week 7, and 3.60 and 3.56 mg/dL at week 23 (Cohen’s d = 0.14, 95%CI: -0.58-0.29) in the tenapanor and control groups, respectively. There was a significant downward trend in serum albumin levels in both groups ( tenapanor: p < 0.0001, control: p < 0.0001), but no significant between-group difference (p = 0.2003).

**Figure 4.**
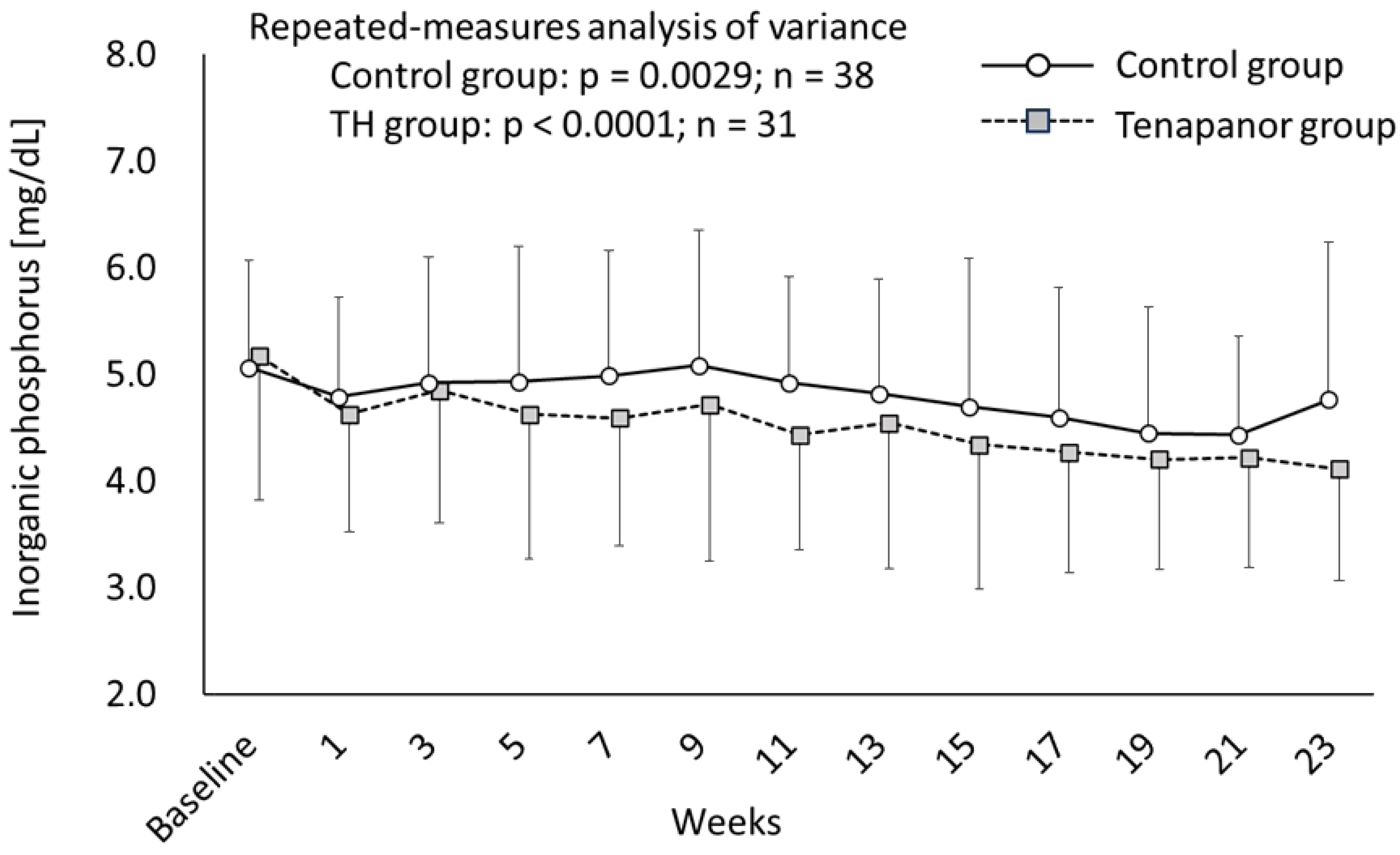
Serum phosphorus level. The data are shown as the mean ± SD.

The Sankey diagram of the changes in BSFS in the tenapanor group is shown in Figure 5. At baseline, 57.3% of patients were considered to have normal stools (types 3 to 5), 25.2% had hard stools (types 1 and 2), and 13.5% had hard stools. During the first week of treatment with tenapanor, 33.3% (n = 6) of patients with types 1 or 2 changed to type 7, and 3 of these patients discontinued treatment with tenapanor by the second week. After tenapanor administration, overall BSFS increased and the proportion of patients with type 6 and 7 stools increased to 32.6%. On the other hand, the proportion of patients with type 1 and 2 stools decreased, reaching 0% by the fifth week of the medication trial. Among the patients who discontinued the study due to diarrhea symptoms or frequent bowel movements, the 2 patients who discontinued at week 1 had a baseline BSFS type 3-4 that increased to type 7 immediately after starting tenapanor. The 4 patients who discontinued between week 1 and week 2 had baseline BSFS type 1-2, but 3 patients had an increase to BSFS type 7 and 1 patient had an increase to BSFS type 5. These patients had a strong aversion to sudden onset of diarrhea (emission of soft or watery stools). Of the 4 patients who discontinued the study after week 3, 2 showed no major changes in their BSFS type, but they felt uncomfortable with the increase in the number of bowel movements, and the remaining 2 patients discontinued the study because they felt a strong aversion to the frequent defecation of soft stools.

**Figure 5.**
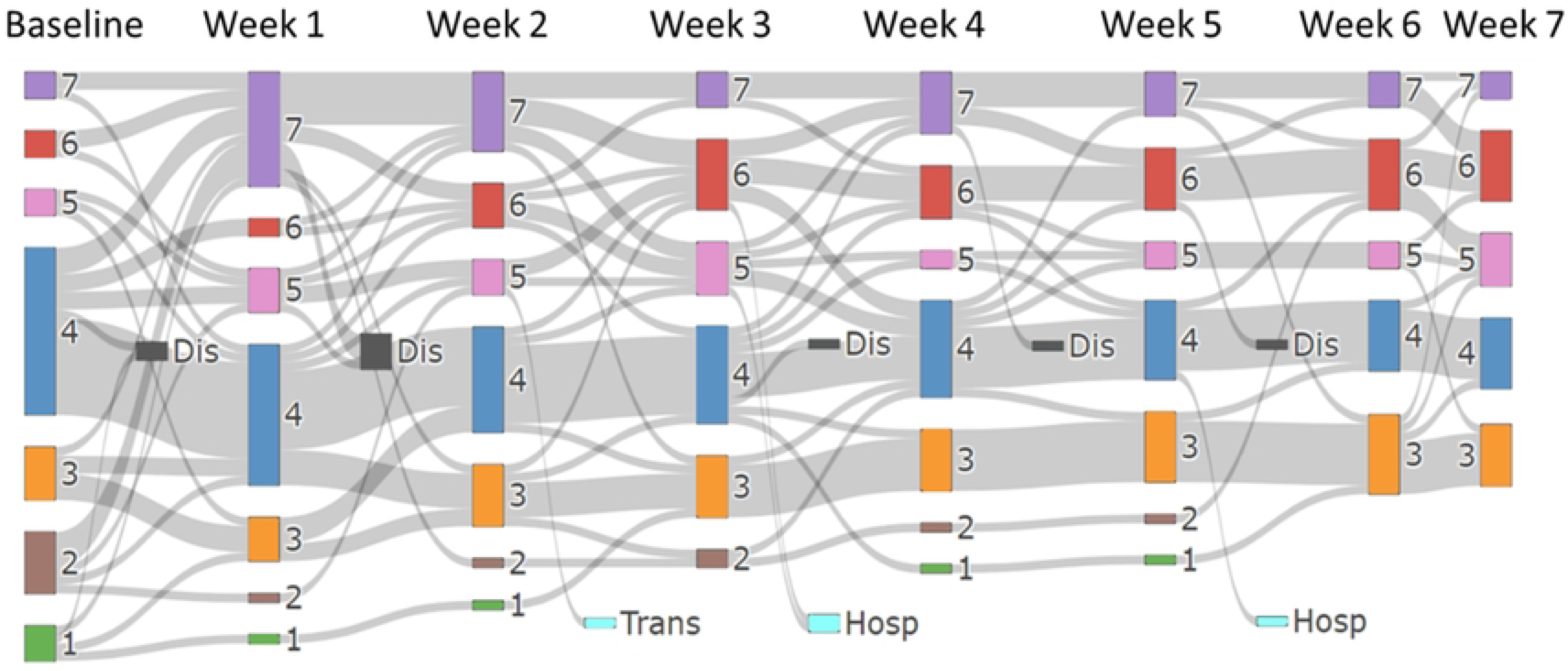
Changes in the Bristol Stool Form Scale in the tenapanor group over seven weeks. Dis; discontinuation, Hosp, hospitalization; Trans, transfer to a different hospital.

The change in the rate of laxative prescriptions is shown in Figure 6. At baseline, 58.2% of patients were prescribed laxatives. At 7 and 23 weeks after starting tenapanor, the proportion of prescription was 21.7% and 35.6%, respectively, a significant decrease from baseline. No subgroup or adjusted analyses were performed for this study as these were not required by the study protocol.

**Figure 6.**
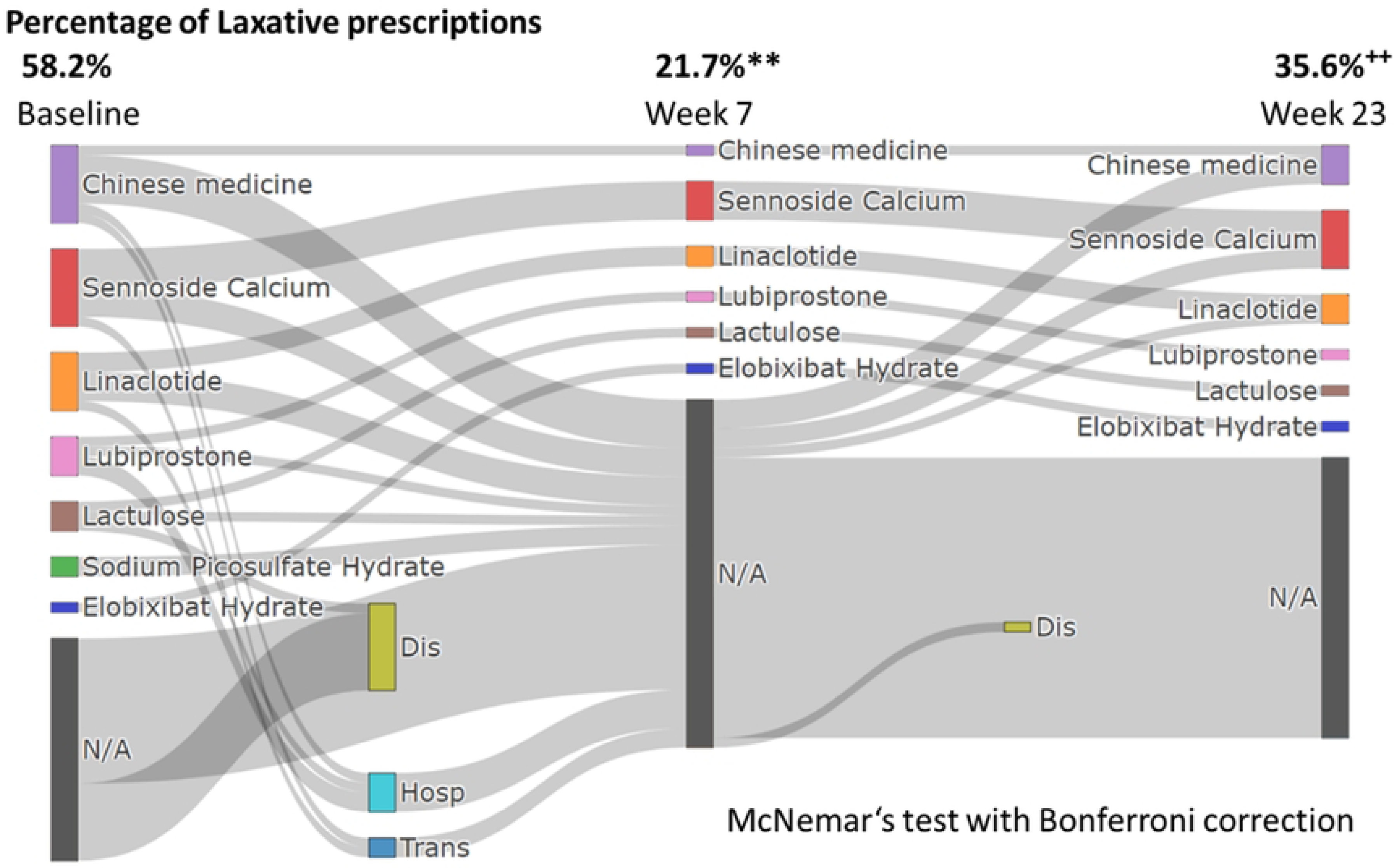
Change in the rate of laxative prescriptions. Dis; discontinuation, Hosp, hospitalization; Trans, transfer to a different hospital. ** indicates a p value less than 0.001 (baseline vs. Week 7). ++ indicates a p value less than 0.001 (Baseline vs. week 23).

## Discussion

The most important finding of this study is that tenapanor improved stool consistency and reduced laxative use in hemodialysis patients while maintaining serum phosphorus control at a low dose, even after switching from lanthanum carbonate. This dual benefit highlights the potential of tenapanor to address both constipation and hyperphosphatemia in clinical practice.

The normalization of stool consistency was notable, with constipation (BSFS types 1-2) completely resolved by week 5. However, a transient increase in loose stools (BSFS types 6-7) was observed early in treatment, consistent with previous studies highlighting diarrhea as a primary adverse effect of tenapanor[10,16-19]. Importantly, the rate of laxative use decreased significantly from 58.2% at baseline to 35.6% at week 23, demonstrating the clinical utility of tenapanor in reducing the medication burden for constipation. However, gastrointestinal intolerance remains a challenge with 10 patients discontinuing treatment due to diarrhea symptoms. In particular, the 6 patients who dropped out of the study by week 2 all showed a rapid increase in BSFS type, and the reason for discontinuation was aversion to this change. This transient increase in diarrhea at the start of tenapanor administration may be due to the rapid excretion of accumulated feces in patients with chronic constipation. A similar phenomenon has been reported in studies of other agents that rapidly improve motility in patients with constipation, where loose stools or diarrhea occur transiently as the bowel evacuates accumulated stool. For example, a study on polyethylene glycol, an osmotic laxative that draws water into the intestines, has shown initial loose stools in the early phase of treatment, thought to be due to the sudden expulsion of impacted stool in constipated individuals[22]. Although direct evidence that tenapanor has this side effect is limited, the pattern observed in this study is consistent with these findings and warrants further investigation.

While lanthanum carbonate is considered the most potent phosphate binder[23], there is a risk of lanthanum deposition in the gastrointestinal tract, which may lead to concerns about long-term safety[12,13]. In this study, the use of low-dose tenapanor effectively suppressed serum phosphorus levels, despite a significant decrease in the prescription of lanthanum carbonate. Furthermore, in two RCTs evaluating the addition of tenapanor to a stable dose of a phosphate binder or placebo, the dual mechanism approach of combining tenapanor with a phosphate binder was shown to significantly reduce serum phosphorus levels[24-25]. The results of these studies highlight the potential of tenapanor to reduce phosphorus levels and suggest that it may serve as a potential replacement for lanthanum carbonate. The proportion of prescriptions for Bixalomer increased during the study period. Bixalomer has been reported to cause constipation as a complication[26], and it is believed that there was a clinical intent to use Bixalomer to correct the tendency for loose stools.

This study has several limitations. Serum albumin levels decreased in both groups. Although the magnitude of the decrease was small, the possibility that it affected phosphorus levels cannot be excluded. The sample size was relatively small, which may have limited the power to detect statistically significant differences, particularly for secondary outcomes. The observation period may have been insufficient to fully evaluate long-term safety and efficacy. In addition, the analyses of secondary outcomes were not adjusted for multiplicity, which increases the risk of false positives. The lack of strict adherence monitoring and the concomitant use of other phosphate binders may have confounded the results. Finally, the open-label design may have introduced bias because both participants and investigators were aware of the group assignments. Future studies with larger cohorts, longer follow-up periods, and stricter treatment protocols are needed to confirm these findings and to evaluate the long-term safety and efficacy of tenapanor.

In conclusion, tenapanor demonstrated significant clinical benefit by improving stool consistency and reducing laxative use while maintaining serum phosphorus control, even after switching from lanthanum carbonate. Individualized treatment strategies may further optimize outcomes. Future studies are needed to explore this hypothesis, clarify long-term safety, and confirm the efficacy of tenapanor in larger populations.

**Supplementary Figure 1. Serum calcium levels corrected for albumin.** The data are shown as the mean ± SD.

**Supplementary Figure 2.** Serum albumin level. The data are shown as the mean ± SD.

## Funding

This study did not receive any specific funding.

## Data Availability

All XXX files are available from the Harvard Dataverse (Link to view the page: https://dataverse.harvard.edu/dataset.xhtml?persistentId=doi:10.7910/DVN/WI6SAS).

## Acknowledgement

None.

## Author Contributions

**Formal analysis:** Naoki Suzuki

**Investigation:** Naoki Suzuki, Yuuki Takeda, Akie Kabuto, Takao Konishi, Takahiro Konishi, Fumi Sumino, Hayanari Iwata, Mone Iwagami, Yusuke Kouti, Yasumasa Hitomi, and Masato Nishimura

**Methodology:** Naoki Suzuki and Masato Nishimura

**Supervision:** Nodoka Sato, Masato Nishimura.

**Writing – original draft:** Naoki Suzuki.

## Notes

### Competing Interest Statement

The authors have declared no competing interest.

### Clinical Trial

the University Hospital Medical Information Network Clinical Trials Registry under trial number UMIN000033778 (UMIN-CTR URL: http://www.umin.ac.jp/ctr/index.htm) We acknowledge that there is a discrepancy between the date of trial registration and the date of participant recruitment. Recruitment for this study was conducted from February 1, 2024, to February 28, 2024, while the trial was registered with the University Hospital Medical Information Network Clinical Trials Registry (UMIN-CTR) on December 25, 2024. This oversight occurred due to an unintentional administrative delay in completing the trial registration process. We recognize the importance of prospective trial registration in ensuring transparency, minimizing potential biases, and aligning with ethical and regulatory guidelines. To address this, we took the following steps: Full Disclosure: We are reporting this discrepancy in the manuscript to maintain transparency. Protocol Consistency: The trial was conducted strictly in accordance with the original protocol, which was developed and finalized before the start of participant recruitment. No changes were made to the study design, eligibility criteria, data collection procedures, or statistical analysis plans after the study was initiated. Ethical Oversight: The study received ethical approval from the Ethics Committee for Human Research at Tojinkai Hospital (Approval Number: 2024-03) prior to the commencement of recruitment. Written informed consent was obtained from all participants before enrollment. We sincerely regret this oversight and have implemented measures to ensure that all future studies are prospectively registered before recruitment begins. We believe that this explanation, combined with the steps taken to ensure the integrity of the study, demonstrates our commitment to upholding ethical standards and transparency in research.

### Funding Statement

The author(s) received no specific funding for this work.

### Author Declarations

Ethical approval was granted by the Ethics Committee for Human Research at Tojinkai Hospital (Approval Number: 2024-03).

